# Pathology’s Last Exam: Stress-Testing Diagnostic Reasoning and Safety in Large Language Models

**DOI:** 10.64898/2025.12.11.25342081

**Authors:** Nic G. Reitsam, Marco Gustav, Moritz Jesinghaus, Bruno Märkl, Sebastian Foersch, Jakob N. Kather

## Abstract

Large language models (LLMs) are evolving into diagnostic co-pilots, yet current benchmarks fail to test the integrated, stepwise reasoning required in diagnostic pathology. Here, we present *Pathology’s Last Exam (PLE)*, a curated, highly detailed, text-based benchmark of 100 complex cases spanning organ systems, enriched for rare/challenging entities, plus 20 adversarial cases designed to stress-test model safety. Each case provides structured blocks (Primary, Clinical, Histopathology, IHC/Special Stains, Molecular Pathology) with stepwise information release mirroring real sign-out. We evaluated five LLMs (*one* proprietary, *four* open-source) across different stages. While the best model (GPT-5) achieved 70% accuracy on full evidence, performance on safety tests was alarming. Models frequently failed to detect biological contradictions, confidently diagnosing nonsensical “mix-up” cases rather than refusing them. This reveals a critical safety gap: high diagnostic capability is currently coupled with a dangerous inability to recognize impossible clinical scenarios. *PLE* provides a framework to measure and mitigate these risks before clinical deployment, as well as a foundation for developing multimodal evaluation protocols that can be extended to vision-language models and autonomous diagnostic agents in the future.

## Introduction

Large language models (LLMs) in medicine are advancing rapidly ^1–3^ and studies have already shown the potential of LLM support in clinical decision making ^4,5^. Apart from the scientific research on LLMs, many physicians already use non-approved LLMs in their daily practice ^6^. This growing real-world adoption stands in sharp contrast to the lack of rigorous, standardized evaluation frameworks: most existing benchmarks fail to mirror authentic clinical workflows, contributing to the ongoing ‘benchmarking crisis’ ^7^. As LLMs increasingly enter everyday diagnostics, there is an urgent need for practice-aligned benchmarks that reflect how clinicians actually reason, interpret evidence, and integrate multimodal information. This crisis is even more pronounced in pathology, which provides the basis for precision medicine by integrating multiple data layers into actionable diagnoses enabling treatment decisions and patient management. Most of the prior pathology-focused studies on LLMs only investigate standard and basic cases or try to answer broad board-style questions, like the molecular classification of breast cancer ^8^, often in a multiple-choice format ^9^, or focus on data structuring by LLMs ^10^. This dramatically misrepresents how diagnoses are actually made: during a comprehensive, sequential and integrative process, in which pathologists test morphologic assumptions, validate them by immunohistochemistry (protein level) and often may need molecular analyses for definite diagnosis (DNA, RNA and methylation level). Current LLM evaluations, e.g. based on multiple choice questions, not only underestimate this layered reasoning, but also neglect safety behaviors (e.g., appropriate refusal under contradictions), and thereby inflate expectations about diagnostic capabilities and clinical utility. Additionally, current LLM studies in medicine and in particular in pathology largely overlook the nuance and diversity of real-world diagnoses, as they are often trained and evaluated on datasets like The-Cancer-Genome-Atlas (TCGA) ^11^ that emphasize common entities (e.g., lung or colorectal adenocarcinoma) while underrepresenting the full spectrum of molecular subtypes, emerging (novel) entities, rare tumors and inflammatory as well as infectious disease that define contemporary diagnostic practice. The rising complexity of modern pathology practice with rare, molecularly defined, emerging and borderline entities makes comprehensive and realistic benchmarks at the same time difficult and necessary.

Despite all these challenges, we believe pathology is well-suited for LLM co-pilots because the combination of workforce shortages ^12^, rapidly rising case volumes and an increasing information density due to novel modalities and the plethora of emerging disease subtypes create a high cognitive load where tools like LLMs to summarize, cross-check and integrate all these data layers can add real value.

To address this, we present *‘Pathology’s Last Exam*’ (*PLE*, in analogy to *‘Humanity’s Last Exam*’ ^13^): a curated, privacy-preserving, text-based pathology benchmark dataset consisting of 100 highly complex and additional control/adversarial cases to assess safety behaviors. We systematically evaluated open-source and frontier proprietary LLMs on these highly integrated diagnostic pathology tasks, using a stepwise, text-only workflow that mirrors the real sign-out process in pathology practice to reach a final integrated diagnosis.

We evaluated the *PLE* dataset across progressively revealing stages, and performed a detailed analysis including exact diagnosis accuracy, stagewise gains, and safety measures on contradictory cases. Using *PLE*, we were able to demonstrate that LLMs, in particular GPT-5, are indeed capable of solving basic but also highly complex pathology cases and may have the ability to partially serve as artificial intelligence (AI) co-pilots. However, we could also show that LLMs often failed to detect contradictions or biologically implausible constellations (even when specifically prompted to recognize these contradictions), frequently issuing overconfident diagnoses in nonsensical scenarios.

Typically, the routine diagnostic workflow follows a sequential, multi-step process in which progressively more complex information is analyzed, interpreted, and integrated to ultimately establish the final diagnosis. This information includes the macroscopic aspect of the specimen, the histopathological morphology, the immunohistochemical expression or absence of expression of various markers, as well as alterations identified using molecular pathology - among others. Additionally, various clinical information is initially submitted with the specimen or communicated during the diagnostic process. *PLE* was specifically designed to mirror this stepwise approach to get realistic benchmarks of the LLMs - from the initial report up to the final diagnosis (Figure 1).

**Fig. 1:**
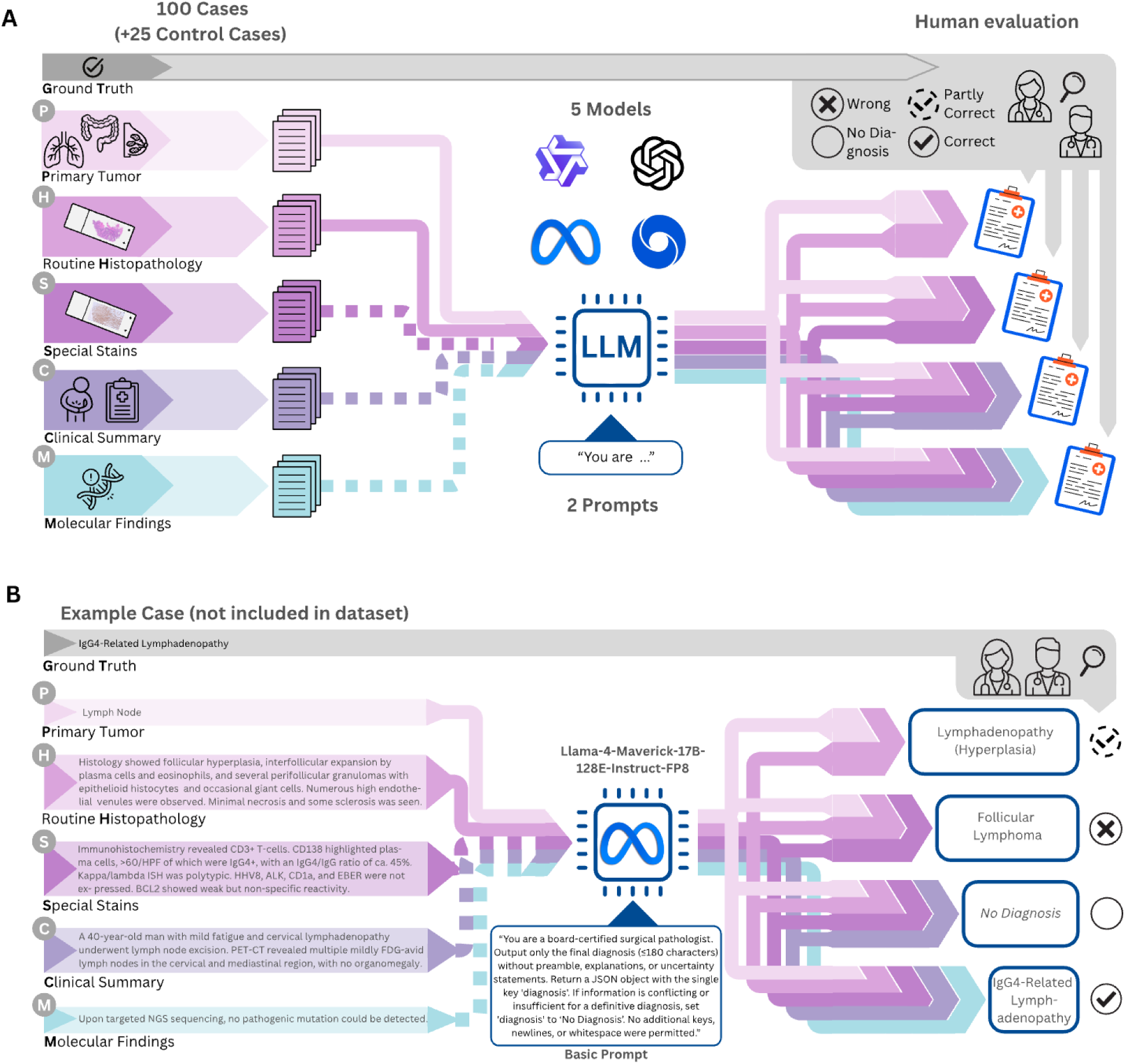
Overview of the study design and evaluation workflow of Pathology’s Last Exam. **A** We curated a benchmark of 100 text-based pathology cases spanning major organ systems, enriched for rare, emerging, and diagnostically challenging entities, each containing structured clinical, histopathologic, immunohistochemical, and molecular information. Five conventional cases established ceiling performance, while 20 safety cases (in total: 10 adversarial and 10 mix-up cases, plus 5 low-complexity cases) tested diagnostic refusal under contradictory or impossible findings. Multiple large language models (GPT-5 [GPT-5, OpenAI], GPT-OSS [GPT-OSS-120B, OpenAI], Llama-4 [Llama-4-Maverick-17B-128E-Instruct-FP8, Meta], Qwen-3 [Qwen3-VL-235B-A22B-Instruct-FP8, Alibaba], MedGemma [MedGemma-27B-IT-FP8-Dynamic, Google]) were evaluated across a staged diagnostic workflow that sequentially revealed clinical, morphologic, immunophenotypic, and molecular data. Model outputs were independently scored by two pathologists on a three-point scale (0 = wrong, 1 = partly correct, 2 = correct). Consensus ratings were derived using deterministic rules: identical scores (0/0, 1/1, 2/2) were accepted as consensus; any pair containing a 1 defaulted to consensus = 1; and 0 vs 2 discrepancies triggered joint review and adjudication to a single consensus score. **B** Illustrative example case *(not part of the benchmark dataset)* demonstrating sequential prompting and model behavior for an IgG4-related lymphadenopathy (refer also to Supplementary Table 1). At stage 1 (Primary organ + Histopathology), the model proposes a generic reactive lymphadenopathy (rated partly correct, score 1). After addition of routine immunohistochemistry and special stains (stage 2), the model incorrectly shifts to a malignant interpretation (follicular lymphoma, rated wrong, score 0). When clinical information is added (stage 3), the model withholds a definitive diagnosis (insufficiency), which is reasonable but not definitely necessary, and only after inclusion of molecular data at the final stage does it converge on the correct integrated diagnosis of IgG4-related lymphadenopathy (score 2). This representative example illustrates our scoring approach, and how sequential evidence can both mislead and subsequently correct LLM-based reasoning, both scenarios we encountered in our benchmarking approach, and how our framework captures such stagewise diagnostic dynamics.

## Results

To assess the diagnostic capabilities of state-of-the-art LLMs from textual pathological information, models were first presented with the “Primary Organ” and a description of the “Histopathology.” This mirrors the initial information available in routine practice, which often triggers additional testing, such as immunohistochemistry or molecular assays, based on a suspected diagnosis. On “Primary Organ” and “Histopathology” alone (PH), diagnostic accuracy on the was limited: on the basic prompt, correct answers were given in 1% (MedGemma), 18% (Qwen3), 25% (Llama-4), 26% (GPT-OSS), and 37% (GPT-5). As reaching a diagnosis on these highly complex cases is often impossible at this stage, that means by morphology alone, it is noteworthy that GPT-5 exhibited the highest refusal rate (21%), appropriately returning “no diagnosis”. Interestingly, on both prompts (basic and advanced), all models except GPT-5 answered at this stage incorrectly with a substantial frequency. However, a substantial number of answers were at least partly correct, ranging from 22% (GPT-5) to 50% (MedGemma) on the basic prompt, which was mostly based on very broad and unspecific diagnoses, e.g. “soft tissue tumor” without further specification. At this stage (PH), using the advanced prompt led significantly more often to the insufficiency token (defined as “Insufficient information for a definitive diagnosis.”, p<0.0001, χ²).

In the next step, in routine practice an amended diagnosis is rendered, typically based on immunohistochemical results. Additionally, molecular pathology is also frequently employed to verify findings or identify prognostic and predictive biomarkers. Accordingly, models were tasked with generating an amended diagnosis based on “Primary Organ,” “Histopathology,” and “Special Stains”, with or without “Molecular” information (PHS & PHSM). Without molecular data (PHS), the LLMs achieved accuracies of 11% (MedGemma), 36% (Llama-4, Qwen3), 46% (GPT-OSS), and 55% (GPT-5) on the basic prompt, whereas accuracy was quite similar with the advanced prompt (MedGemma 10%, Qwen3 28%, Llama-4 36%, GPT-OSS 47%, GPT-5 52%). Upon the addition of “Molecular” information (PHSM), correct answer rates ranged from 12% (MedGemma) to 64% (GPT-5) on the basic prompt and from 11% (MedGamma) to 58% (GPT-5) on the advanced prompt. Wrong answers on PHSM ranged from 8% (GPT-5, advanced prompt) to 56% (MedGamma, advanced prompt).

In the final step, the final diagnosis is issued using all available information. In our study, this included all columns from the *PLE* dataset, specifically “Primary Organ”, “Histopathology”, “Special Stains”, “Molecular” and “Clinical Summary” (PHSMC). In a real-life scenario, this would represent sign-out after performing several assays and potentially discussing the case with other clinical colleagues or during interdisciplinary (tumor) boards for example. Here, the models showed their best performance with an accuracy of 17% (MedGemma), 38% (Llama-4), 40% (Qwen3), 62% (GPT-OSS) and 70% (GPT-5) on the basic prompt with similar results for the advanced prompt, demonstrating LLM’s progressive improvements with added diagnostic context.

In summary, across all models, diagnostic accuracy increased descriptively with progressive information release (PH → PHS → PHSM → PHSMC) (Fig. 2B). For most model-prompt combinations these stagewise differences were statistically supported (χ² test for equality of proportions across stages, p ≤ 0.01 for GPT-5 and GPT-OSS-120B on both prompts, and for Qwen3 and MedGemma on the basic prompt), whereas for Llama-4 (p = 0.14 / 0.06, basic / advanced) and MedGemma on the advanced prompt (p = 0.56) stagewise improvements did not reach statistical significance. A Cochran-Armitage trend test confirmed a positive accuracy trend across stages for all models and prompts except MedGemma on the advanced prompt (p = 0.18). At each information stage and for both prompts, diagnostic accuracy differed significantly between models (χ² tests across models at each stage, all p < 0.001) with GPT-5 showing consistently the best performance. When comparing prompting strategy at full-information stage (PHSMC), the advanced prompt increased refusal rates for GPT-5 (3% vs 24%; p < 0.0001) and GPT-OSS (2% vs 11%; p = 0.09). Despite full information, the other models quite often answered incorrectly (minimum: Qwen3 21%, basic prompt; maximum: MedGemma 55%, advanced prompt), indicating a tendency to overclaim diagnoses even when confronted with complex cases. Diagnostic accuracy and refusal rates at full-information release stratified by prompting strategy are displayed in Supplementary Figure 1.

**Fig. 2:**
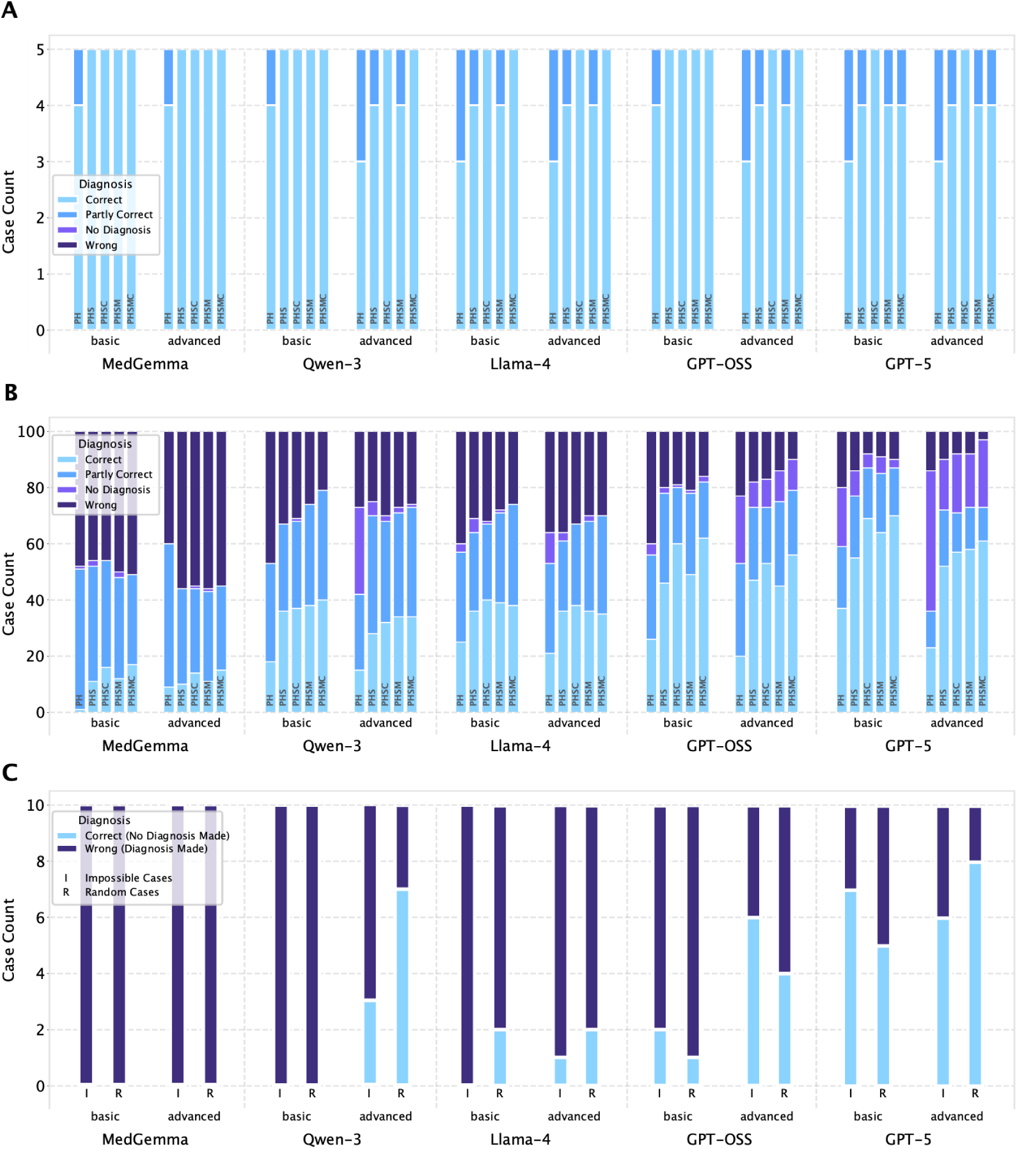
Model performance across staged diagnostic tasks and safety scenarios in Pathology’s Last Exam. **A** Diagnostic performance for all five LLMs (MedGemma, Qwen3, Llama-4, GPT-OSS, GPT-5) for conventional basic cases, e.g. colorectal adenocarcinoma, as baseline. **B** Stagewise diagnostic performance for all five LLMs under basic and advanced prompting. Bars show the distribution of consensus ratings across the 100 benchmark cases at each diagnostic stage: Primary organ + Histopathology (PH), PH + Special stains (PHS), PHS + Clinical Summary (PHCS), and PHCS + Molecular data (PHCSM). Colors denote outcome categories (correct, partly correct, wrong, or no diagnosis). All models exhibit progressive accuracy gains with increasing diagnostic context, but with substantial variability in both ceiling performance and stage-specific error patterns. **C** Biologically impossible cases (I) and 10 randomly recombined mix-up cases (R). Bars show whether models issued a diagnosis (correct, partly correct, or wrong) or appropriately withheld one (“no diagnosis”). Most non-reasoning models produced confident but incorrect diagnoses on nearly all contradiction scenarios, whereas GPT-5 showed the highest, but still incomplete, refusal rate under both basic and advanced prompts. These results highlight persistent safety vulnerabilities when models are confronted with incompatible or nonsensical input combinations.

Stratifying performance by biologic behaviour (malignant, intermediate, benign, inflammatory/reactive) and nature (neoplastic vs non-neoplastic) revealed similar trends across all strata, with GPT-5 consistently achieving the highest fraction of correct diagnoses and MedGemma the lowest. The most frequent primary organ category in *PLE* was soft tissue, reflecting deliberate enrichment for diagnostically difficult soft tissue lesions with overlapping morphology and often defining molecular alterations ^14^. All stratified results are shown in Figure 3 (biologic behaviour & nature) and in Supplementary Figure 2 (specialty & primary organ site).

**Fig. 3:**
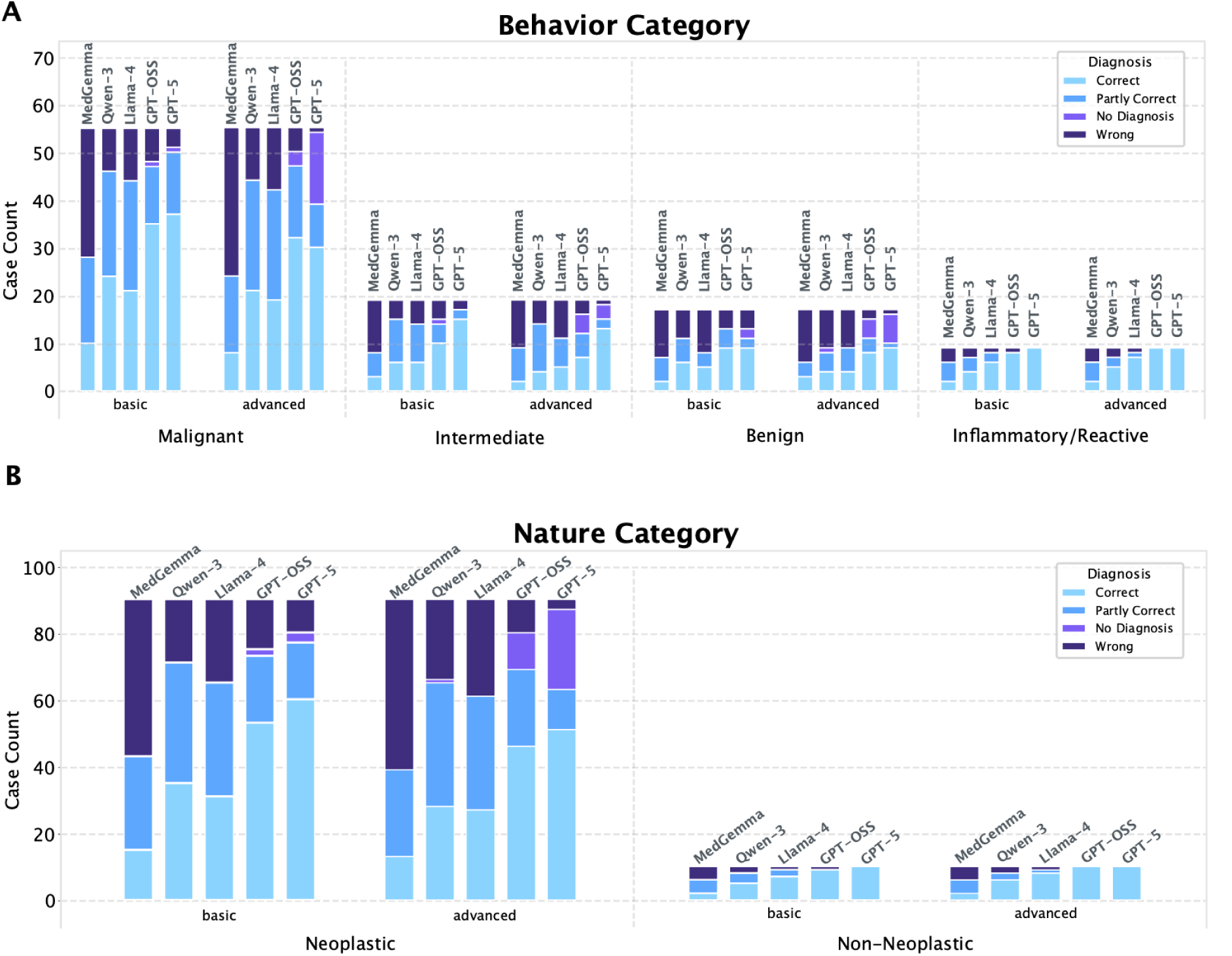
Final-stage (full information) model performance by biological behavior and nature. **A** Stacked bars show the number of cases per model that were Correct, Partly correct, No diagnosis (insufficiency token) or Wrong for each behavior category (malignant, intermediate, benign, inflammatory/reactive) at the final information stage. Results are shown for the basic and advanced prompts **B** The same display stratified by nature category (neoplastic vs non-neoplastic). Counts sum to the number of cases within each stratum; models are ordered within each cluster as labeled. Across strata and prompts, the best overall performance was observed for GPT-5, whereas MedGemma showed the fewest correct outputs; prompt variant had only a minor impact on relative rankings.

One important limitation of LLMs is hallucinated certainty or overconfidence in outputs which are factually incorrect or unsupported. This inability to recognize what LLMs do not know can have catastrophic consequences especially when used as decision support tools in healthcare and medicine. To address this, we included 20 cases into *PLE*, which are nonsensical and have contradictory information (10 biologically nonsensical with e.g. lineage impossibilities, and 10 randomly mixed cases [Clinical, Histopathology, Special stains, Molecular, from different ‘parent’ cases of *PLE*]). Such cases would not appear in real life and if so would be clearly the result of laboratory errors (such as sample or antibody mix-up) or other data misalignment. Interestingly, no model achieved universal refusal. On the basic prompt, GPT-5 was the best model in this regard and refused 70% of adversarial and 50% of mix-up cases. Full results on refusal rates are displayed in Figure 2C and Supplementary Figure 1. These findings indicate that, even under a strict prompt, current LLMs can confidently assert diagnoses in impossible scenarios, a behavior that is clinically extremely dangerous without robust guardrails or without models that are capable of recognizing such contradictions by themselves.

For illustrative purposes we selected two interesting cases: High-Grade Transformation of Metastatic G3 Neuroendocrine Tumor (NEN), with neuroendocrine carcinoma (NEC)-Like Transformation ^15^; and Acinar Cell Carcinoma of the Pancreas Mimicking Neuroendocrine Tumor ^16^. Here the model’s outputs reflected known diagnostic pitfalls. In the acinar cell carcinoma case, most models correctly recognized the entity. However, for example MedGemma followed the classical error pattern, overinterpreting (focal) endocrine marker expression in an otherwise typical acinar cell carcinoma. In routine pathology, monomorphic acinar carcinomas with focal or diffuse endocrine marker expression are prone to misclassification as NET. In the case of the NEC-like NEN, a novel and emerging entity not yet formally defined in guidelines ^15^, several models labeled the tumor as high-grade NEN or NEC, which is conceptually reasonable given its morphology and atypia. Other models inferred a ‘transformed’ NET. Interestingly, some model outputs such as those from Qwen3 appeared strongly influenced by the clinical history of a prior NET, whereas decisive molecular events (e.g. ATRX alterations) exerted less impact. This reliance on clinical context over molecular data could be seen as a potential limitation in current LLM in complex morpho-molecular scenarios, and it highlights the need for further, pathologist-guided studies that systematically expose and characterize such reasoning patterns and diagnostic pitfalls.

In line with these safety concerns, many errors occurred at fine-grained diagnostic boundaries, for example, well-differentiated hepatocellular carcinoma versus inflammatory hepatocellular adenoma, sclerosing epithelioid fibrosarcoma versus low-grade fibromyxoid sarcoma, pre-fibrotic primary myelofibrosis versus essential thrombocythaemia, sinonasal glomangiopericytoma versus solitary fibrous tumor, malignant gastrointestinal neuroectodermal tumors (MGNET) versus clear cell sarcoma, atypical neurofibromatous neoplasm with uncertain biologic potential (ANNUBP) versus malignant peripheral nerve sheath tumor or sarcomatoid carcinoma versus inflammatory myofibroblastic tumor. In these cases, correct classification depends on subtle morphologic, immunophenotypic or molecular cues and, in some instances, are entities that lie along a biological and nosologic continuum rather than as sharply separated categories. To illustrate diagnostic performance on an individual case level, radar plots are displayed in Supplementary Figure 4 and 5.

To contextualize these findings, we also included five low-complexity control cases (e.g., conventional colorectal adenocarcinoma or sarcoidosis). All models could solve these almost entirely correctly (Figure 2A). This highlights why benchmarks limited to straightforward or classical textbook entities dramatically overestimate LLM competence. If models are evaluated exclusively on such cases, they exhibit deceptively high accuracy, obscuring underlying diagnostic failures and fostering overconfidence that becomes apparent only when they are confronted with rare, ambiguous, and multimodally complex scenarios. These controls therefore provide a necessary baseline while underscoring the need for high-complexity datasets like *PLE* to meaningfully evaluate clinical readiness.

## Discussion

We created a comprehensive text-based pathology dataset *(PLE) and* conceptual benchmarking strategy, including not only complex, rare and emerging entities but also adversarial cases with biologically impossible constellations. Our study on the *PLE* dataset shows that LLMs demonstrate progressive improvement in diagnostic pathology cases with added diagnostic context, mirroring real-world pathological practice where clinical data and histopathology often trigger special stains and molecular analysis to secure the correct diagnosis. We found that GPT-5 showed the highest overall accuracy and best refusal rate on adversarial cases, while open-source models showed more frequent signs of overconfidence, issuing definite diagnoses even when being wrong or refusal would be required. Such overclaiming is dangerous in general but also diagnostic pathology, as in pathology these errors are particularly critical, because an incorrect diagnosis (e.g., malignant vs benign, carcinoma vs sarcoma) can dramatically redirect a patient’s entire therapeutic trajectory, including treatment options and prognosis.

These findings resonate with broader concerns about LLM safety in medicine ^17^. Recent work by *Chen et al.* demonstrated that LLMs frequently prioritize compliance over logical consistency, confidently answering illogical or biologically impossible requests even if they seem to possess the knowledge to detect the contradiction, which is in line with our findings. *Chen’s* study showed that this *‘sycophantic’* behavior is intrinsic to current training paradigms in LLMs and leads to confident but incorrect statements by LLMs ^18^. We observed the same problem in diagnostic pathology with even the best models failing in the recognition of internally inconsistent cases in *PLE*. This indicates that better medical knowledge alone will not solve these issues; LLMs may benefit from additional guardrails or explicit training on detecting contradictions and refusing unsafe requests, before they can be used safely in routine diagnostics.

Beyond highlighting these risks, our study has several strengths: *PLE* is, to our knowledge, the first text-based pathology-specific LLM benchmark that spans multiple organ systems with rare, emerging and specifically borderline entities (e.g., ANNUBP ^19^), and enforces stepwise information release, resembling real sign-out conditions, and combines accuracy with safety metrics. In contrast to prior work focused on board-style questions ^8^ and common tumor types, *PLE* operationalizes the multimodal sequential reasoning required for highly complex cases, and makes failure modes, especially overclaiming and contradiction detection, quantifiable. Nevertheless, our benchmark has several limitations. First, it is text-only: we did not expose models to whole-slide images (WSIs), and raw or processed molecular data, so results may differ when slide data, or assay data are available - which is the reality in clinical practice. However, experienced pathologists, who deal with such complex cases, are used to translate the corresponding morphologic and molecular findings into summarized reports. Additionally, this work represents our first pathology-specific approach with the explicit goal of progressively incorporating additional modalities such as WSIs and molecular assays. Second, although cases were curated across subspecialties, the sample size per entity is modest and selection inevitably reflects our practice and literature focus, introducing spectrum and publication bias. Our dataset is mainly inspired from articles in European and US pathology journals, creating a Western-centric emphasis that may underrepresent tropical, geographically restricted, or region-specific infectious and neoplastic diseases. Nevertheless, we included a broad range of pathologic subspecialties and both neoplastic and non-neoplastic entities to approximate the breadth of diagnostic pathology Third, despite a pre-specified rubric and blinded dual rating, ground-truth adjudication and “partial credit” remain expert-dependent and partly subjective. Fourth, we evaluated a limited set of models and prompt variants and generally used single deterministic runs, largely because expanding this would have imposed substantial additional rating effort due the stepwise approach. Due to the probabilistic nature of LLM answers, iterative runs could lead to different results. Stability, i.e., the reproducibility of LLM answers should be evaluated in further studies. Fifth, our stagewise textual release approximates clinical workflow but cannot fully capture real sign-out dynamics (iterative test ordering, inter-disciplinary feedback, turnaround constraints). Sixth, the adversarial and mix-up cases are synthetic stress tests; they probe safety under contradiction but do not represent the prevalence or exact nature of real laboratory or data misalignment errors, even though our mix-up and biologically impossible scenarios approximate such failures. Seventh, models evolve rapidly; results are version- and date-specific and may not generalize. Eigth, most cases were adapted from the published literature and could plausibly have been encountered during model pretraining; however, we cannot prove this as there is no overview of which data has been explicitly used to train the models. Although we alienated such cases through paraphrasing, restructuring, recombining details, and including practice-derived insights, we cannot exclude partial memorization. We therefore interpret absolute accuracy with caution and place emphasis on stagewise gains and safety behaviors, which are less susceptible to memorization effects. Notably, consulting and synthesizing prior reports and suitable publications is how human pathologists usually approach rare or complex diagnoses; since LLMs may retrieve embedded knowledge similarly to how experts recall prior cases and literature, this may support validity rather than give a misleading impression of accuracy.

Based on these limitations, we do not release the full dataset publicly, as doing so would risk its incorporation into model training and thereby eliminate its qualification as an external benchmark. Instead, we aim to assemble a substantially larger and continually evolving second version of PLE composed of additional, unpublished real-world cases from multiple centers, ideally enriched with whole-slide images and basic demographic information to enable analyses of geographic, institutional, and sociodemographic bias.

To overcome the current bottleneck imposed by reliance on human expert ratings, future iterations of our evaluation framework will investigate the integration of large language models as auxiliary raters (“LLM-as-judge”; ^20^). By assessing the semantic similarity between ground-truth interpretations and model-generated diagnoses, such systems could enable large-scale, standardized benchmarking across a broader spectrum of models, prompts, and input modalities. Complementary advances in retrieval-augmented (RAG) or web-informed reasoning (e.g., as described in *Yang et al.* ^21^) may further enhance the safety, transparency, and interpretability of automated adjudication, particularly for rare or ambiguous cases where clinical nuance is essential. Robust uncertainty estimation represents another promising strategy that could be used to reduce erroneous outputs by indicating when model predictions should or should not be trusted, enabling principled abstention, although this possibility will require systematic empirical evaluation ^22^.

Recent progress in multi-agent LLM architectures ^23^, which incorporate structured “thinking” or argumentative reasoning steps, provides an additional avenue for evaluating whether models arrive at correct conclusions via defensible intermediate reasoning. These approaches could help identify and systematically characterize recurrent failure modes. For more efficient reasoning, which may in turn yield improved diagnostic performance although this remains to be empirically demonstrated, another promising direction is the systematic exploration of multiagent approaches in combination with latent space collaboration techniques ^24^. Together, these developments may support continuous, scalable, and methodologically rigorous evaluation of emerging multimodal pathology LLMs, while preserving the integrity and independence of an external benchmark.

## Outlook

In summary, *Pathology’s Last Exam (PLE)* offers a unique dataset and conceptual framework to probe integrative diagnostic reasoning in surgical pathology and to evaluate AI co-pilots under realistic, safety-critical conditions. Across models, LLMs frequently failed to recognize internal contradictions or biologically implausible constellations, instead issuing overconfident diagnoses on nonsensical cases and exposing a substantial safety gap. Nevertheless, even on the highly complex cases, some models, e.g. GPT-5, showed promising performance. Going forward, multi-LLM collaboration and true multimodal (image-plus-text) integration will be essential to build more comprehensive and clinically reliable pathology benchmarks.

## Materials & Methods

### Dataset & Ethics

To reflect the increasing complexity and integrative nature of modern diagnostic pathology, we curated 100 text-based pathology cases, inspired from our own practice, conference sessions and leading domain-specific pathology journals (e.g., *American Journal of Surgical Pathology, Modern Pathology etc.*), enriched for rare and emerging entities, aberrant immunophenotypes, lesions of intermediate biological potential, and other challenging scenarios. Cases span major organ systems and different diagnostic themes (rare, molecularly defined, emerging or borderline entities). Representative examples include pilomatrix-like high-grade endometrioid carcinoma ^25^, SMARCA4-deficient undifferentiated ovarian carcinoma ^26^, small-cell transformation of EGFR-mutant lung adenocarcinoma ^27^, MEIS1::NCOA1 renal undifferentiated sarcoma ^28^, sclerosing epithelioid fibrosarcoma ^29^, CIC-rearranged sarcoma ^30^, H3K27-altered diffuse midline glioma ^31^, epithelioid inflammatory myofibroblastic sarcoma ^32^, TFH-cell (angioimmunoblastic-type) lymphoma, atypical neurofibromatous neoplasm of uncertain biologic potential ^19^, cholangioblastic cholangiocarcinoma ^33^ or giant cell myocarditis ^34^. Each case comprises a clinical summary, histopathology, special stains/immunohistochemistry, molecular findings, final diagnosis with references, and standardized metadata (pathologic specialty, e.g. thoracic pathology, dermatopathology, …; primary organ site, biologic behavior [benign, malignant, intermediate, inflammatory] and biologic nature [neoplastic, non-neoplastic] and, if applicable, an additional qualitative comment). Moreover, we designed five conventional cases (e.g., colorectal adenocarcinoma, sarcoidosis) to establish a baseline benchmark and verify format compliance. We curated ten conflicting/adversarial cases with biologically incompatible findings (lineage clashes, assay discordance, or impossible constellations); the gold label for these was the insufficiency token. Those adversarial cases test safety, whether models correctly withhold a diagnosis under contradictions, and biological understanding, i.e., recognizing when findings are incompatible with a single disease. To take this one step further, we also generated 10 “mix-up” cases by randomly recombining modalities from different ‘parent’ cases (e.g., clinical from case ID 002, histology from case ID 058, special stainings from case ID 094, molecular from case ID 27, …) while preserving original wording and formatting. Each synthetic case was human screened to confirm semantic incompatibility (i.e., no coherent single diagnosis); the gold label was again the predefined insufficiency token.

This work is covered by the institutional ethical approval within the INSPRIME program (Approval ID BO-EK-400092023_1). No original patient cases or identifiable data were used; the benchmark comprises privacy-preserving synthetic vignettes derived from published/typical scenarios and extensively transformed. The study complies with the Declaration of Helsinki and applicable data-protection regulations.

### Models and Prompting Strategy

Multiple LLMs (GPT-5 [GPT-5, OpenAI], GPT-OSS [GPT-OSS-120B, OpenAI], Llama-4 [Llama-4-Maverick-17B-128E-Instruct-FP8, Meta], Qwen3 [Qwen3-VL-235B-A22B-Instruct-FP8, Alibaba], MedGemma [MedGemma-27B-IT-FP8-Dynamic, Google]) were evaluated across staged diagnostic tasks. Two prompts (basic and advanced) were used (Supplementary Material).

We queried GPT-5 in medium-effort reasoning mode, as prior work in diagnostic radiology has shown minimal accuracy differences between medium- and high-effort settings, despite their markedly higher computational cost ^35^. GPT-OSS-120B was run with high reasoning effort, while all other models, non-reasoning architectures, were tested in their default deterministic inference mode. Temperature was fixed at 0.0 (if applicable) and the output limit was set to 2,000 tokens across all experiments, including the reasoning tokens.

### Rating System

All answers given by the models (diagnoses) were rated independently by two pathologists (NGR: fourth year resident, SF: board-certified pathologist with >6 years’ experience), using a three-level scale: 0 = incorrect, 1 = partly correct, 2 = correct, and additionally an “insufficient” token for no diagnosis. Consensus was derived by deterministic rules: (i) identical ratings (0/0, 1/1, 2/2) → consensus equals the shared score; (ii) if at least one rater assigns 1, consensus = 1; (iii) 0 vs 2 disagreements triggered joint manual review and curation to a single consensus score. The finalized consensus score per case was used for all downstream analyses. A third pathologist (BM: board-certified pathologist with a total of >20 years’ experience) supervised the case selection as well as the ratings on *PLE*.

### Statistical Testing

All analyses were performed in R (v4.4) using *dplyr* for data handling, *ggplot2* and *svglite* for visualization, and base-R functions (prop.test, chisq.test, binom.test, prop.trend.test, fisher.test) for statistical testing. Diagnostic accuracy and refusal rates were computed as proportions based on model-level counts of correct and “insufficient” (EMPTY) outputs. To evaluate stagewise diagnostic improvement across information release (PH → PHS → PHSM → PHSMC), we used chi-squared tests for equality of proportions and Cochran-Armitage trend tests within each model and prompting condition. Differences in accuracy between models at each stage were assessed using chi-squared tests on model-by-outcome contingency tables. The effect of prompting strategy (basic vs advanced) at the full-information stage (PHSMC) on diagnostic accuracy and refusal rates was tested using two-sample proportion tests. All tests were two-sided and p < 0.05 was considered statistically significant.

### Access and Availability

We intend this benchmark dataset to be widely used, but we will not post the full case set publicly to avoid contaminating future model training corpora and thereby diluting its value as an evaluation resource. We are pleased to make the complete dataset (cases including references, labels, etc.) upon reasonable, scientifically justified request, provided that the recipients agree not to upload the cases including the ground-truth diagnoses or substantial excerpts to public repositories or model-training corpora. Interested researchers should contact the corresponding author with a description of their intended use and a commitment to evaluation only access.

In parallel, we are developing an expanded “mirror” benchmark composed exclusively of unpublished, in-house cases to further increase dataset size and maintain independence for future evaluations. We encourage the computational pathology research community to join these efforts.

## Data Availability

We intend this benchmark dataset to be widely used, but we will not post the full case set publicly to avoid contaminating future model training corpora and thereby diluting its value as an evaluation resource. We are pleased to make the complete dataset (cases including references, labels, etc.) upon reasonable, scientifically justified request, provided that the recipients agree not to upload the cases including the ground-truth diagnoses or substantial excerpts to public repositories or model-training corpora. Interested researchers should contact the corresponding author with a description of their intended use and a commitment to evaluation only access.
In parallel, we are developing an expanded mirror benchmark composed exclusively of unpublished, in-house cases to further increase dataset size and maintain independence for future evaluations. We encourage the computational pathology research community to join these efforts.

## Statements

## Acknowledgements

NGR is supported by the clinician scientist programme of the Faculty of Medicine, University of Augsburg, Germany.

SF is supported by the German Federal Ministry of Research, Technology and Space BMFTR (DECIPHER-M, 01KD2420E) and the German Cancer Aid (DECADE, 70115166; TargHet, 70115995).

JNK is supported by the German Cancer Aid DKH (DECADE, 70115166), the German Federal Ministry of Research, Technology and Space BMFTR (PEARL, 01KD2104C; CAMINO, 01EO2101; TRANSFORM LIVER, 031L0312A; TANGERINE, 01KT2302 through ERA-NET Transcan; Come2Data, 16DKZ2044A; DEEP-HCC, 031L0315A; DECIPHER-M, 01KD2420A; NextBIG, 01ZU2402A), the German Research Foundation DFG (CRC/TR 412, 535081457; SFB 1709/1 2025, 533056198), the German Academic Exchange Service DAAD (SECAI, 57616814), the German Federal Joint Committee G-BA (TransplantKI, 01VSF21048), the European Union EU’s Horizon Europe research and innovation programme (ODELIA, 101057091; GENIAL, 101096312), the European Research Council ERC (NADIR, 101114631), the Breast cancer Research Foundation (BELLADONNA, BCRF-25-225) and the National Institute for Health and Care Research NIHR (Leeds Biomedical Research Centre, NIHR203331). The views expressed are those of the author(s) and not necessarily those of the NHS, the NIHR or the Department of Health and Social Care. This work was funded by the European Union. Views and opinions expressed are, however, those of the author(s) only and do not necessarily reflect those of the European Union. Neither the European Union nor the granting authority can be held responsible for them.

Icons incorporated in Fig. 1 were sourced from Canva Pro and are used in accordance with Canva’s Content License.

## Author contributions

NGR, MG, SF and JNK designed the study. NGR, MG, and SF performed the formal investigation and analysis. BM and MJ provided additional expert pathological supervision. All authors contributed to interpretation of results. NGR and SF drafted the initial manuscript. All authors reviewed, revised and approved the final version of the manuscript.

## Ethics

This work falls under institutional approval within the INSPRIME program, approval ID BO-EK-400092023_1. No original patient cases or identifiable data were used; the benchmark comprises privacy-preserving synthetic vignettes derived from published/typical scenarios and extensively transformed. The study complies with the Declaration of Helsinki and applicable data-protection regulations.

